# Visual Exploratory Data Analysis of COVID-19 Pandemic: One Year After the Outbreak

**DOI:** 10.1101/2021.05.04.21256635

**Authors:** Amril Nazir, Suleyman Ulusoy, Lujaini Lotfi

**Affiliations:** Department of Information Systems, College of Technological Innovation, Zayed University, Abu Dhabi, United Arab Emirates; Department of Mathematics and Natural Sciences, American University of Ras Al Khaimah, P.O. Box 10021, Ras Al Khaimah, United Arab Emirates; AnalytiCray Solutions, No 2-16, Jalan Pandan Prima 2, 55100, Kuala Lumpur, Malaysia

**Keywords:** COVID-19, visual exploratory data analysis, government interventions

## Abstract

**Background:** Governments across the globe have taken different measures to handle the Covid-19 outbreak since it began in early 2020. Countries implemented various policies and restrictive measures to prevent transmission of the virus, reduce the impacts of the outbreak (i.e., individual, social, and economic), and provide effective control measures. Although it has been over one year since the outbreak started, few studies have examined the long-term effects of the pandemic. Furthermore, researchers need to examine which government intervention variables are the most, and least, effective. Such analysis is critical to determine the best practices in support of policy decisions.

**Methods:** Visual exploratory data analysis (V-EDA) offers a user-friendly data visualization model to evaluate the impact of the pandemic. It allows one to observe visual patterns of trends. The V-EDA was conducted on one year data for the COVID-19 Pandemic, one year after the outbreak between 1 January and 31 December, 2020. The data were analyzed using the student’s t-test to verify if there was a statistical difference between two independent groups, and the Spearman test was also used to analyze the correlation coefficient between two quantitative datasets and their positive or negative inclination.

**Findings:** We found that high-testing countries had more cases per million than low-testing countries. For low-testing countries, however, there was a positive correlation between the testing level and the number of cases per million. This suggests that high-testing countries tested in a preventive manner while low-testing countries may have a higher number of cases than those confirmed. The poorest developing countries have reduced testing which can coincide with the reduction in new cases, which we did not observe in the high-testing countries. Among the restrictive measures analyzed, a higher population aged 70 or older and lower GDP per capita was related to a higher case fatality ratio. Restrictive measures reduce the number of new cases after four weeks, indicating the minimum time required for the measures to have a positive effect. Finally, public event cancellation, international travel control, school closing, contact tracing, and facial coverings were the most important measures to reduce virus spread. We observed that countries with the lowest number of cases had a higher stringency index.

## 1 Introduction

Governments across the globe have taken different measures to handle the Covid-19 outbreak since it began in early 2020. Countries implemented various policies and restrictive measures to prevent transmission of the virus, reduce impacts of the outbreak (i.e., individual, social, and economic), and provide effective control measures. Although it has been over one year since the outbreak started, few studies have examined the long-term effects of the pandemic. Furthermore, researchers need to determine which government intervention variables are the most, and least, effective. Such analysis is critical to determine the best practices in support of policy decisions.

Visual exploratory data analysis (V-EDA) offers a user-friendly data visualization model that evaluates the most effective government approaches for handling the outbreak. [1] was the first to conduct the V-EDA on COVID-19 for China and worldwide using a one month dataset between January and February, 2020. [2] also explored V-EDA to analyze effective measures the Kerala government in India enforced. [3] conducted the most recent work on V-EDA for COVID-19, offering basic preliminary worldwide analysis on the number of positive, recovered, death cases, mortality and recovery rates using the Johns Hopkins University dataset for a 6 month period (i.e., between 22 January and 12 June, 2020).

Existing COVID-19 studies using the V-EDA approach provide important visualization insights, but they lack statistical evidence. They also focus specifically on gaining insights on short-term effects on the government intervention measures, as opposed to long-term effects. Researchers currently have limited knowledge of the long-term effectiveness of various government intervention strategies. Specifically, what are the long-term effects of relaxed restrictive measures versus extreme ones on particular government intervention? Furthermore, policymakers and governments often need to make a well-informed decision on whether to enforce moderate or extreme measures on a specific intervention. What is the guideline for such a decision to be made?

We conducted a V-EDA of the global COVID-19 pandemic one year after the outbreak between 1 January and 31 December, 2020, to better understand the long-term effects of certain government policies. Additionally, we aimed to measure to the efficacy of such policies. Our analysis focuses on the effects of relaxed versus extreme restrictive measures on a particular government intervention or policy implementation.

We also examined the effects of moderate restrictive measures (at 75th percentile of the population) versus extreme measures (at 95th percentile) with the aim to gain additional insight into the success of the policy implementation based on the strictness level. Policymakers may then make decisions to enforce moderate or extreme measures based on the evidence. Statistical evidence supports our V-EDA, validating the visual observations and preventing biased insights. Our results offer important insights into the best practices and the effective responses that governments and policymakers can implement for successful COVID-19 control.

We analyzed the public datasets using the V-EDA approach with statistical reporting to examine the following questions:

- What is the effect on the number of positive cases when countries conduct high or low numbers of tests?
- Do countries that conduct the most tests have better outcomes in containing the virus? What is the effect of conducting more tests?
- What is the effect of conducting few tests?
- Can a country reduce Case Fatality Ratio (CFR) by conducting more tests as an early precautionary measure to prevent patients from developing health complications that lead to death?
- Does the CFR strongly correlate with countries’ hospital bed capacity, countries’ economic output, the elderly population, and the median population age?
- Do the strictest measures have a better chance to control the outbreak? What is the impact on positive cases if countries enforce strict or lenient lockdown policies?
- What is the effect of implementing aggressive lockdown and restrictive measures? Does it significantly help reduce cases?
- What will be the ramifications if countries have lenient public lockdown measures? Does it have negative consequences?
- For countries that effectively controlled the COVID-19 outbreak, which government interventions (i.e., staying at home, internal movement restriction, travel control restrictions, etc.) had the most success? What are important predictor variables that can prevent or minimize the spread of COVID-19?
- What are the consequences of containment measures on the COVID-19 outbreak?
- Do countries with the strictest lockdown polices have advantages in containing the virus when compared to countries with lenient lockdown policies?

To the best of our knowledge, this is the first work that performs V-EDA on COVID-19 in this comprehensive evidence-based manner. We aim to answer the above questions using data for a one year period.

## 2 Materials and Methods

### Data

We conducted a V-EDA of the global COVID-19 pandemic based on the one year public datasets between 1 January and 31 December, 2020. We assembled the data from two main sources which are publicly available: the Our World in Data Research group (OWID)—a scientific online publication that focuses on large global problems–and the Oxford COVID-19 Government Response Tracker (OxCGRT)—research work from an academic team at the University of Oxford. Both datasets are structured in tabular format as Comma-Separated Values format (CSV).

The OWID dataset comprises 52 variables which provides daily assessment of the number of positive cases, death cases, and tests. OWID also consists of variables on the number of hospital beds, the country’s GDP per capita, the country’s median age, and the distribution of the elderly population (aged 70 or older). We selected 6 variables from the OWID dataset to generate the visual exploratory graphs for Figure 1 to Figure 5: location, date, new daily cases, new daily cases per million, new daily tests per thousand, and new daily deaths. Table 1 provides a summary of selected features the OWID dataset uses.

**Table 1:** Selected features used for OWID Dataset

| Variable Name | Description |
| --- | --- |
| location | The name of country |
| date | The date of day |
| new_tests_per_thousand | The number of new tests per thousand of day |
| new_cases | The number of new cases of day |
| new_cases_per_million | The number of new cases per million of day |
| new_deaths | The number of new deaths of day |

**Figure 1:**
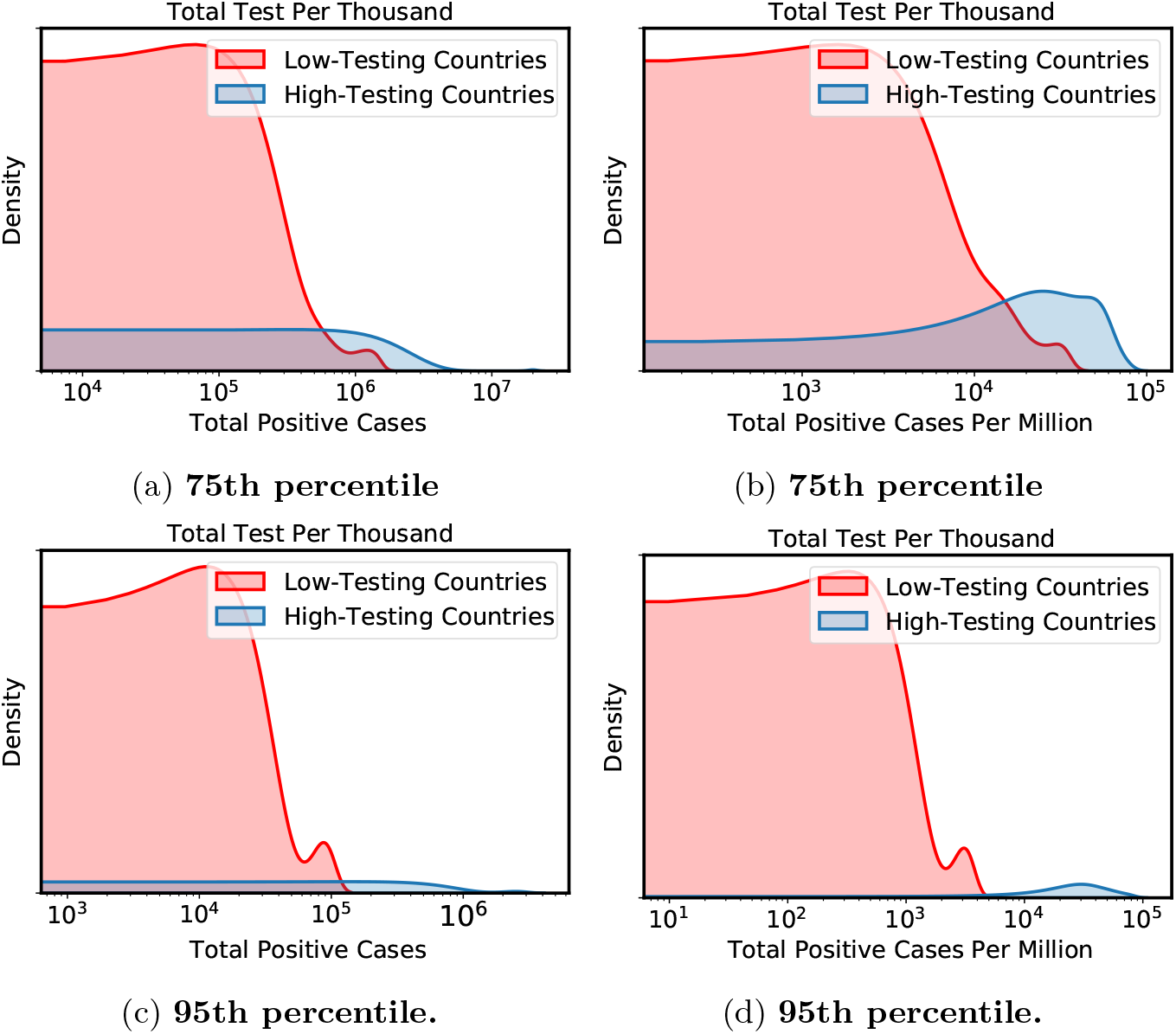
What is the effect on the number of positive cases when countries conduct high or low numbers of tests? Let’s assume *x*_0_ as an arbitrary value on the x-axis. The probability of having the total positive cases equal to *x*_0_ is the area under the corresponding curve to the left of *x*_0_. Clearly, the area to the left of *x*_0_ under the red curve is larger almost every time with the exception of a few points on the right side of the range for the 75th percentile. This indicates that low-testing countries have higher tendencies to have fewer positive cases. Further, analysis of the difference between low-testing and high-testing countries in A) total positive cases in the 75th percentile of the countries (P=0.13, Student’s t-test). B) total positive cases per million in the 75th percentile of the countries (P*<*0.0001, Student’s t-test). C) total positive cases in the 95th percentile of the countries (P=0.18, Student’s t-test). D) total positive cases per million in the 95th percentile of the countries (P*<*0.0001, Student’s t-test). In low-testing countries, the number of tests is positively correlated with the number of cases per million (r=0.69, P*<*0.0001, Spearman Test). In high-testing countries, the correlation was not significant (r=0.16, P=0.33, Spearman Test).

For the OxCGRT dataset, we analyzed 2 major categories: government responses to COVID-19 (identified as the stringency index), and the related COVID-19 policies on health containment (identified as the health containment index). For the stringency index, we considered 8 variables, including school closing, workplace closing, public event cancellation, restrictions on gatherings, public transport closing, staying at home, internal movement restriction, and international travel control. For the containment health index, we considered 7 variables, including public information campaigns, testing policy, contact tracking, emergency investment in health care, investment in vaccines, facial coverings, and vaccination policy. Table 2 provides a summary of selected features the OxCGRT dataset uses.

**Table 2:**
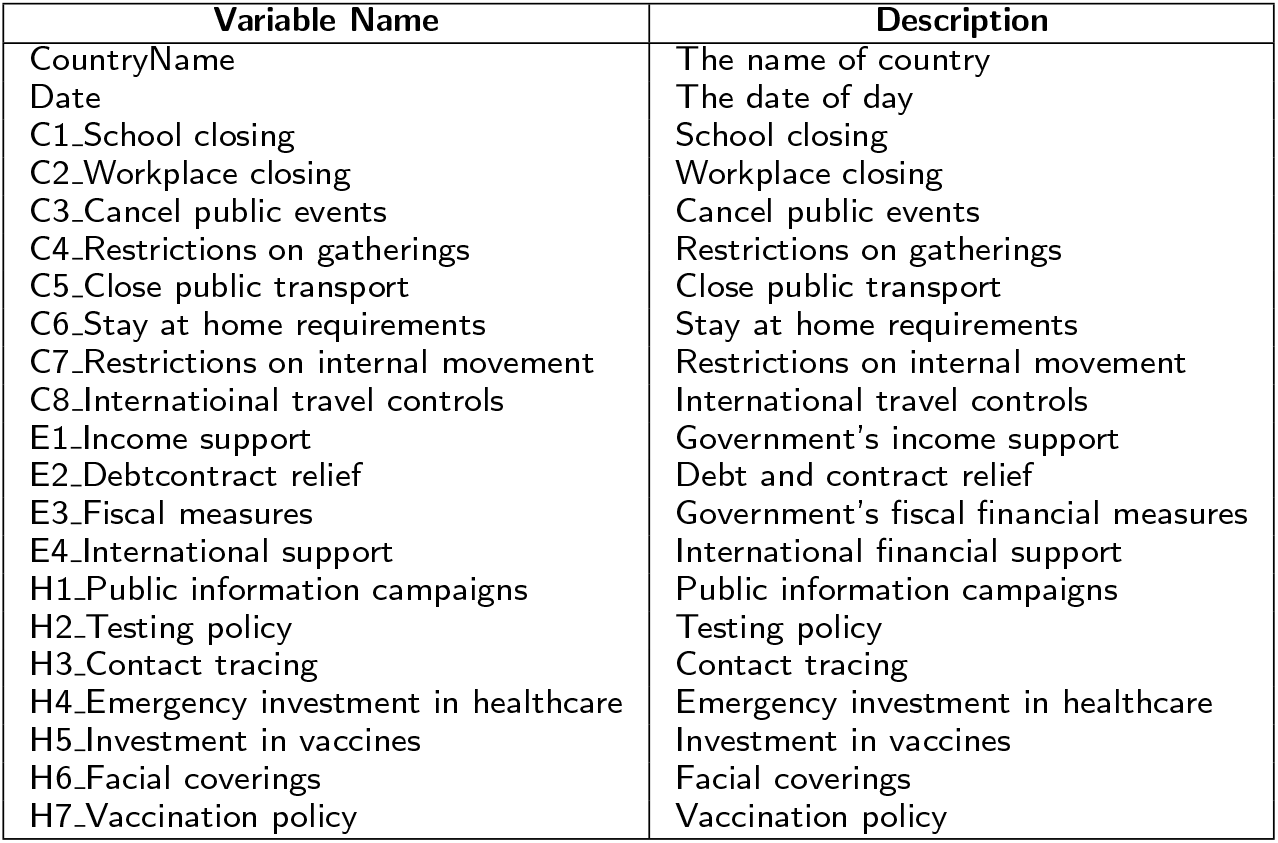
Selected features used for OXCGRT Dataset

### Preprocessing and Visualization

The original OWID dataset covers 191 countries. OWID, however, removed 10 countries because they did not have any data prior to March, 2020. These countries include Comoros, Lesotho, Marshall Islands, Samoa, Sao Tome and Principe, Solomon Islands, South Sudan, Tajikistan, Vanuatu, and Yemen. Therefore, we selected 181 countries for analysis from the OWID dataset.

We observed irregularities in some of the countries’ reported tests and positive case numbers. We identified 76 countries that reported no testing despite reporting positive cases. For example, Venezuela reported no tests despite having reported 113,558 positive cases. We also discovered 1 country (i.e., Hong Kong) with no recorded cases. We removed Hong Kong from our analysis. The original OxCGRT dataset covers 184 countries. We merged the OWID and OxCGRT datasets using the country and the date as the primary keys.

After merging the OWID and OxCGRT datasets, we selected 163 countries for analysis. We removed 18 countries from the OWID dataset and 21 countries from the OxCGRT dataset, since they did not match based on the country names.

We suspected the actual number of cases would be higher for some poorer countries than the number of reported cases due to limited testing, reporting lags, and under-reporting. For Figure 1 and 4, we divided countries into two groups: low-testing and high-testing. We used the commonly accepted threshold of the 5th percentile of the total test per thousand to represent low-testing countries and the 95th percentile of the total test per thousand to represent high-testing countries. To observe the impact of moderate measures (versus extreme measures), we compared our results with the 25th percentile of the countries to represent low-testing countries and the 75th percentile to represent high-testing countries.

For Figure 2 and 3, we normalized the total test per thousand and the total positive case variables based on the monthly percentage change. The trend on the effect of testing is often only visible after a substantial period. Therefore, we make a direct comparison between these two variables on a monthly basis (instead of a daily or weekly basis), in order to identify any relationship. We calculated the monthly percentage C as follows:

**Figure 2:**
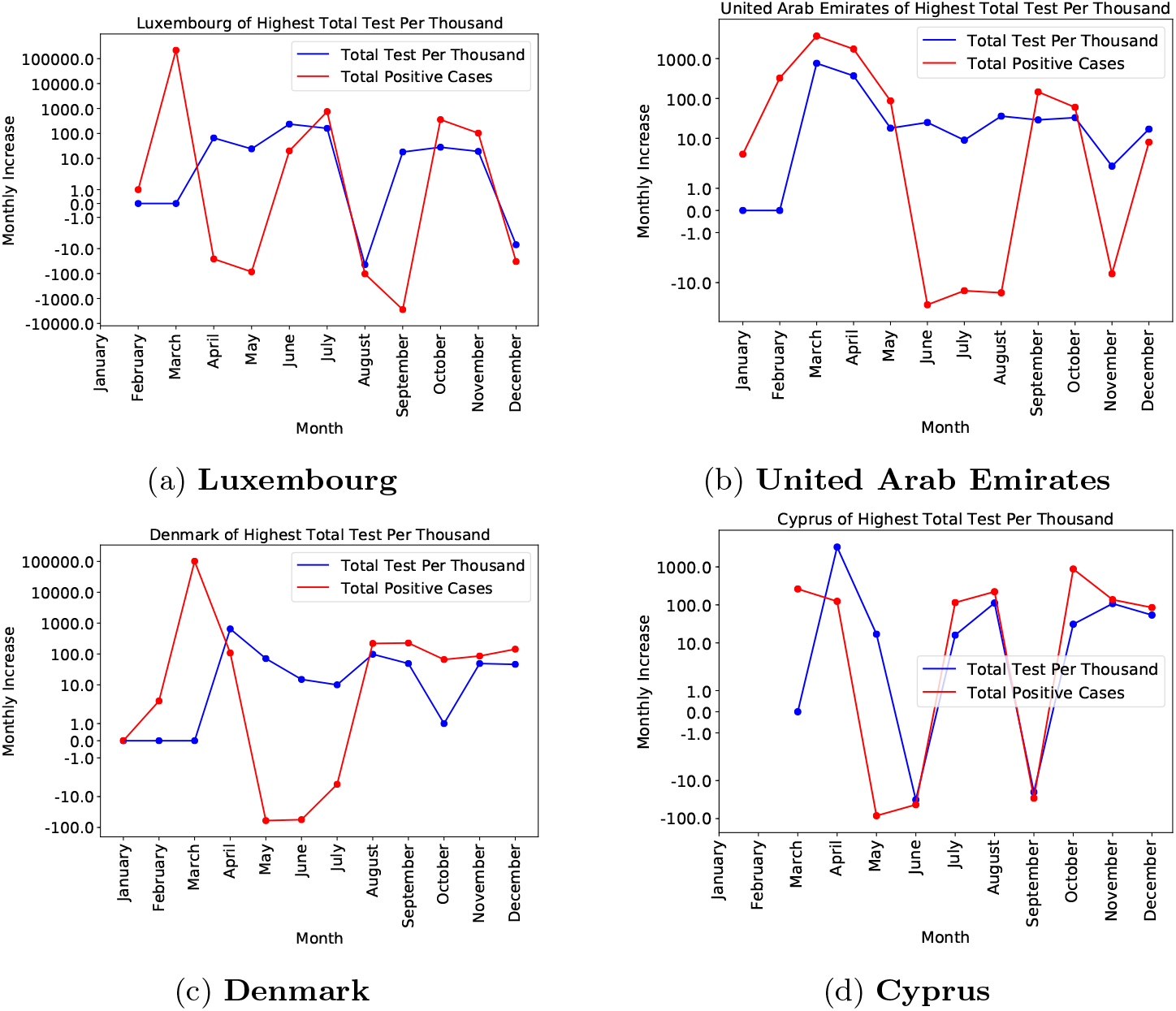
Do countries that conduct the most number of tests have better outcomes in containing the virus? What is the effect of conducting more tests? Total test per thousand and total positive cases by month in the top 4 high-testing countries A) Luxembourg (r=0.38, P=0.25), B) United Arab Emirates (r=0.31, P=0.32), C) Denmark (r=0.13, P=0.69) and D) Cyprus (r=0.41, P=0.25). The data were analyzed using the Spearman Test.

**Figure 3:**
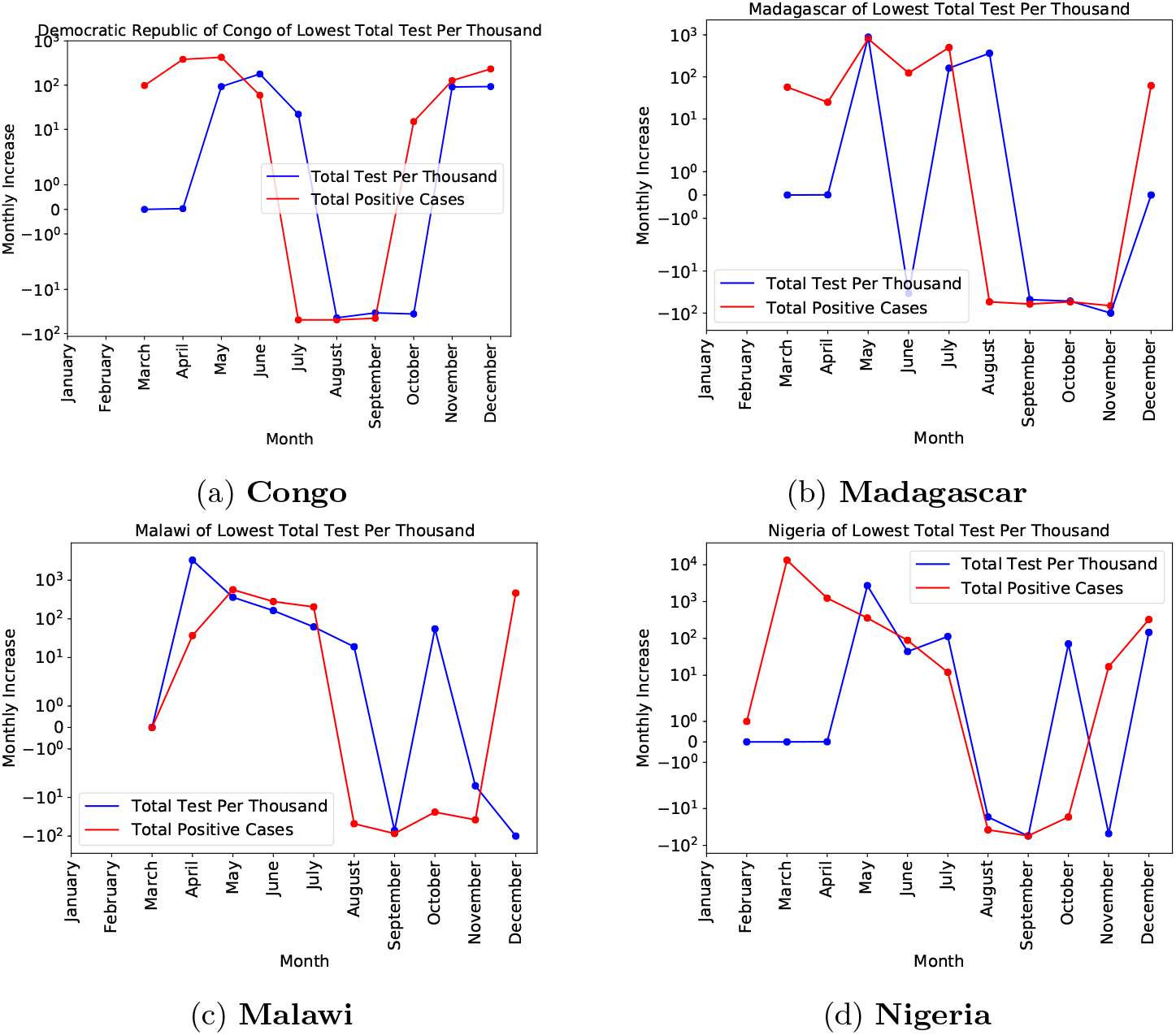
On the other hand, what is the effect of conducting few tests? Total test per thousand and total positive cases by month in the top 4 low-testing countries A) Democratic Republic of Congo (r=0.56, P=0.1), B) Madagascar, positive correlated (r=0.65, P= 0.045), C) Malawi (r=0.42, P-0.23), D) Nigeria (r=0.43, P=0.18). The data were analyzed using the Spearman Test.

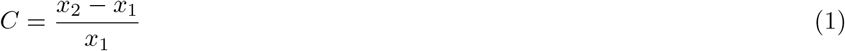

where *x*_1_ is the initial value (i.e., the current month) and *x*_2_ is the final value (i.e., the next month).

For Figure 4, we computed the Case Fatality Ratio (CFR) by dividing the number of daily death cases by the number of daily positive cases. For Figure 5, we used a similar method to separate the two different groups. The 5th and 25th percentiles represent a category of countries with the lowest hospital beds per thousand, GDP per capita, aged 70 or older, and median age. Conversely, the 75th and 95th percentiles represent a category of countries with the highest hospital beds per thousand, GDP per capita, aged 70 or older, and median age.

**Figure 4:**
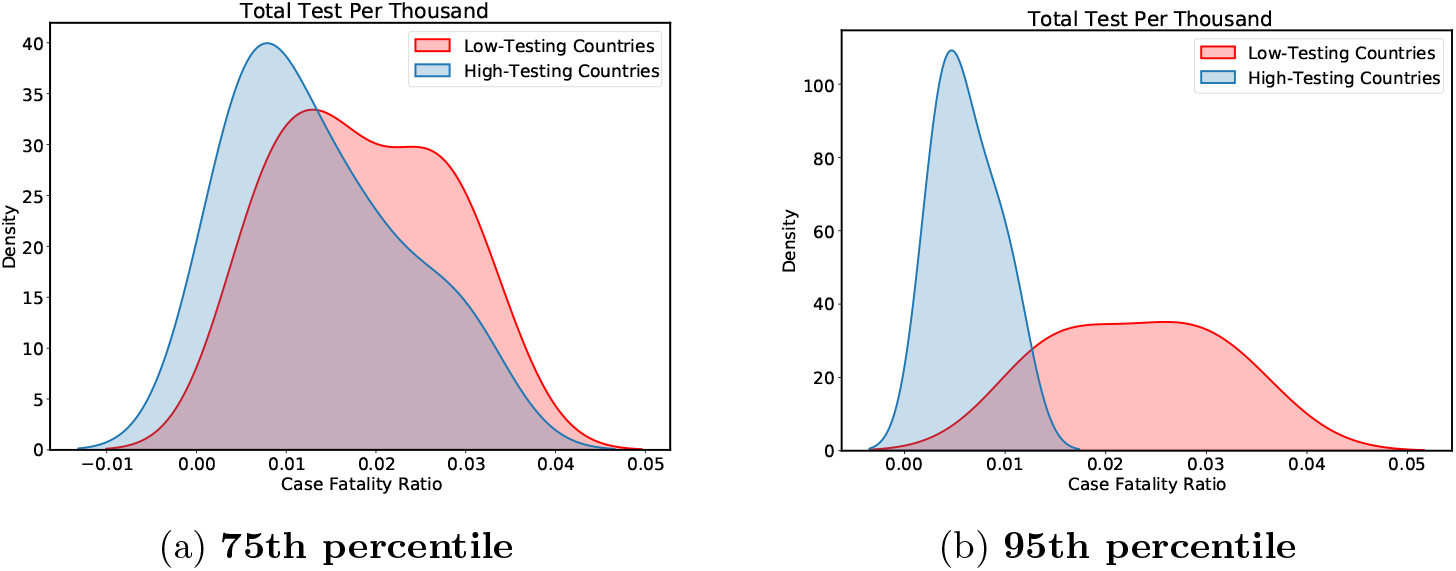
Can a country reduce Case Fatality Ratio (CFR) by conducting a higher number of tests as an early precautionary measure to prevent patients from developing health complications that lead to deaths? We can see that the concentration of the data for the CFR of the high-testing countries is smaller compared to the low-testing countries. From both 75th and 95th percentiles, there is also clear evidence that the higher range of values on the CFR (x-axis) is smaller for high-testing countries indicating that high-testing countries have lower CFR when compared to low-testing countries. Using the Student’s t-test, analysis of the Case Fatality Ratio (CFR) in low-testing and high-testing countries is the following: A) in the 75th percentile, CFR was not different B) in the 95th percentile, low-testing countries had a higher CFR (mean = 0.0231) than the CFR of high-testing countries (mean = 0.0062, P = 0.003).

**Figure 5:**
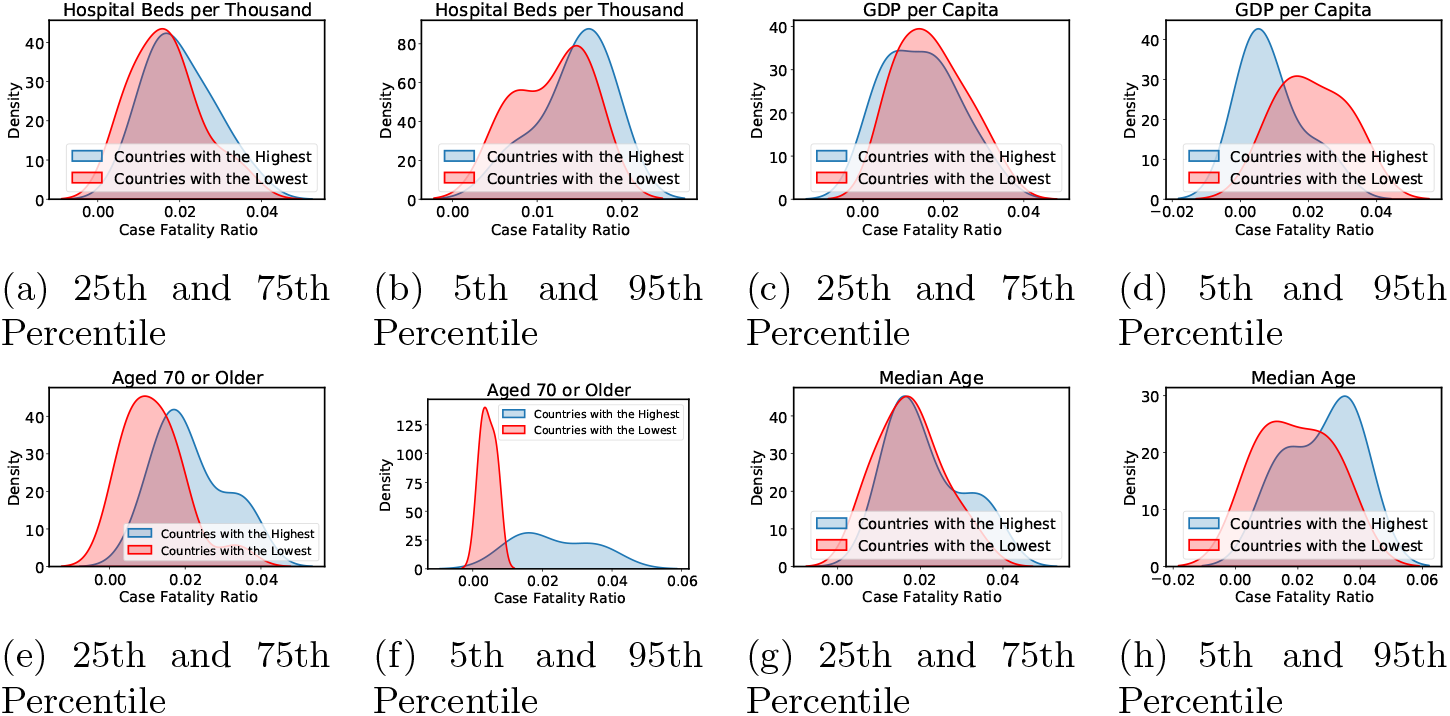
Does the Case Fatality Ratio (CFR) strongly correlate with countries’ hospital bed capacity, countries’ economic output, the elderly population, and the median population age? We can clearly see that population aged 70 was highly related to the CFR. The GDP per capita also seems to have moderate correlation with CFR. Analysis of case fatality ratio related to A) Hospital Beds per Thousand (HBPT) in the 75th percentile countries. The mean HBPT was 8.1 ± 0.6 in the countries with the highest HBPT and was 0.64 ± 0.07 in the countries with the lowest HBPT (P*<*0.0001; 95% confidence interval [CI], −8.8 to −6.2). B) HBPT in the 95th percentile countries. The mean HBPT was 10.6 ± 1.0 in the countries with the highest HBPT and was ± 0.06 in the countries with the lowest HBPT (P*<*0.0001; 95% confidence interval [CI], −12.5 to −8.0). C) GDP per capita in the 75th percentile countries. The mean GDP per capita was 60738 ± 5329 in the countries with the highest GDP per capita and was 2113 ± 243.9 in the countries with the lowest GDP per capita (P*<*0.0001; 95% confidence interval [CI], −69552 to −47698). D) GDP per capita in the 95th percentile countries. The mean GDP per capita was 82275 ± 10182 in the countries with the highest GDP per capita and was 1177 ± 115.3 in the countries with the lowest GDP per capita (P*<*0.0001; 95% confidence interval [CI], −104579 to −57616). E) Percentage of the population aged 70 or older in the 75th percentile countries. The mean population aged 70 or older was 14.1 ± 0.4 in the countries with the highest percentage and was 1.4 ± 0.1 in the countries with the lowest percentage (P*<*0.0001; 95% confidence interval [CI], −13.5 to −11.9). F) Percentage of the population aged 70 or older in the 95th percentile countries. The mean population aged 70 or older was 15.7 ± 0.8 in the countries with the highest percentage and was 1.0 ± 0.2 in the countries with the lowest percentage (P*<*0.0001; 95% confidence interval [CI], −16.5 to −12.8). G) Percentage of the population median age in the 75th percentile countries. The mean population age was 44.7 ± 0.5 in the countries with the highest percentage and was 18.7 ± 0.3 in the countries with the lowest percentage (P*<*0.0001; 95% confidence interval [CI], −27.0 to −24.8). H) Percentage of the population median age in the 95th percentile countries. The mean population age was 46.5 ± 0.7 in the countries with the highest percentage and was 17.4 ± 0.3 in the countries with the lowest percentage (P*<*0.0001; 95% confidence interval [CI], −30.8 to −27.3). The data were analyzed using the Student’s t-test.

The stringency index rates the stringency of government measures to COVID-19 from 0 to 100. The index assigns 100 to countries with the strictest rules or highest lockdown measures. To establish the effects of government interventions on the number of positive cases, we computed the monthly percentage change of total cases for two groups: low and high stringency (Figure 6). The 5th and 25th percentiles represent low-stringency countries and the 75th and 95th percentiles represent high-stringency countries.

**Figure 6:**
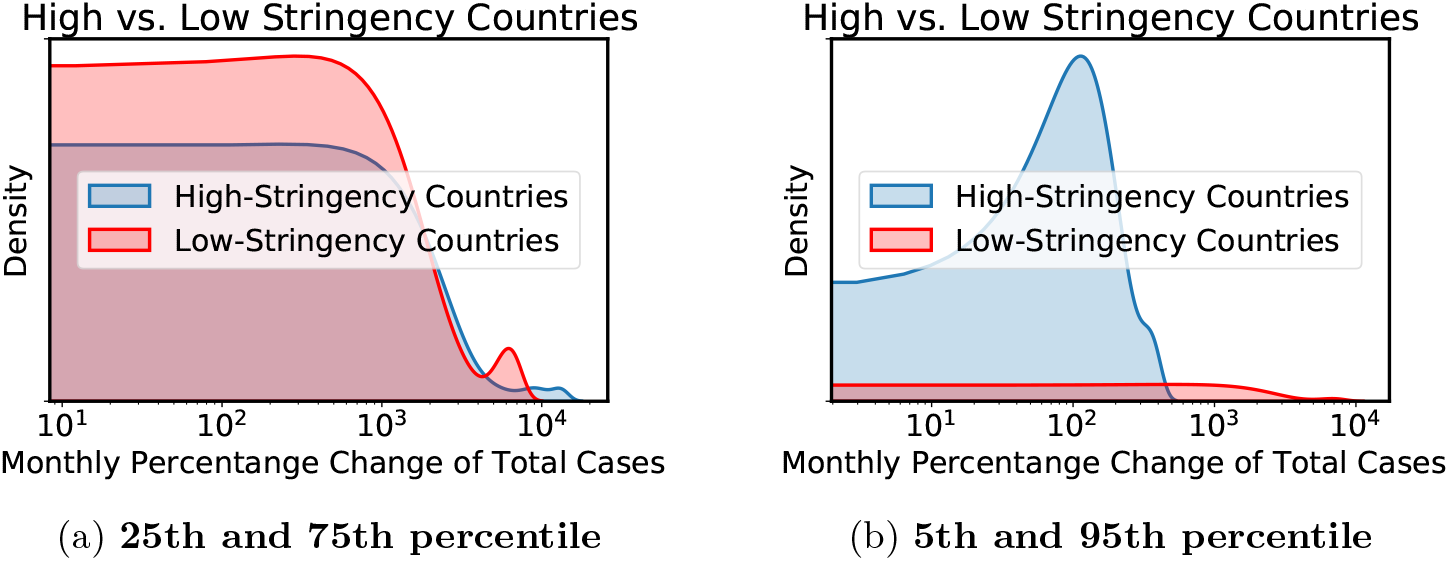
Do the strictest measures have a better chance to control the outbreak? What is the impact on positive cases if countries enforce strict or lenient lockdown policies? Let’s assume *x*_0_ as an arbitrary value on the x-axis. The probability of having the total positive cases equal to *x*_0_ is the area under the corresponding curve to the left of *x*_0_. At 25th and 75th percentile, the area to the left of *x*_0_ under the red curve is larger almost every time with the exception of a few points on the right side of the range. However, an opposite trend is observed at the 5th and 95th percentile whereby the area to the left under the blue curve has significant concentration. This indicates that extreme lockdown measures are essential to have significant reduction on the monthly percentage change of cases. Moderate lockdown measures do not have significant impact on the monthly reduction since both high and low stringency countries have similar concentrations and peaks, and we also observed a few countries at the right side of the range that incurred very high monthly percentage change of total cases under moderate lockdown. Using the Student’s t-test, monthly percentage change of total cases in A) high and low stringency countries in the 75th percentile. The mean monthly percentage change of total cases was 66.2 ± 0.7 in the high-stringency countries and was 31.4 ± 1.8 in the low-stringency countries (P*<*0.0001; 95% confidence interval [CI], −38.6 to −31.0). A) high and low stringency countries in the 95th percentile. The mean monthly percentage change of total cases was 70.3 ± 1.2 in the high-stringency countries and was 21.4 ± 2.8 in the low-stringency countries (P*<*0.0001; 95% confidence interval [CI], −55.4 to −42.5).

Additionally, we computed the monthly percentage change of the total cases to determine the effect of high-stringency policies on the number of cases. To determine the efficacy of high-stringency policies, we calculated the percentage change of total cases for the current month and the following month (Figure 7). If high stringency government policies have a positive impact on the decline of COVID-19 cases, we would expect the monthly percentage change to decrease. Moreover, we computed the monthly percentage change of the total cases of the current month and the following month for low-stringency countries (Figure 8). These calculations examine the effects of countries that have lenient approaches towards lockdown measures. We also extracted all variables from the government stringency and health containment indexes to draw the boxplots (Figure 9 and 10). For these plots, we compute the restrictive measure score by normalizing each value of the variable by its maximum value. The normalized score gives the range of values between 0 and 1.

**Figure 7:**
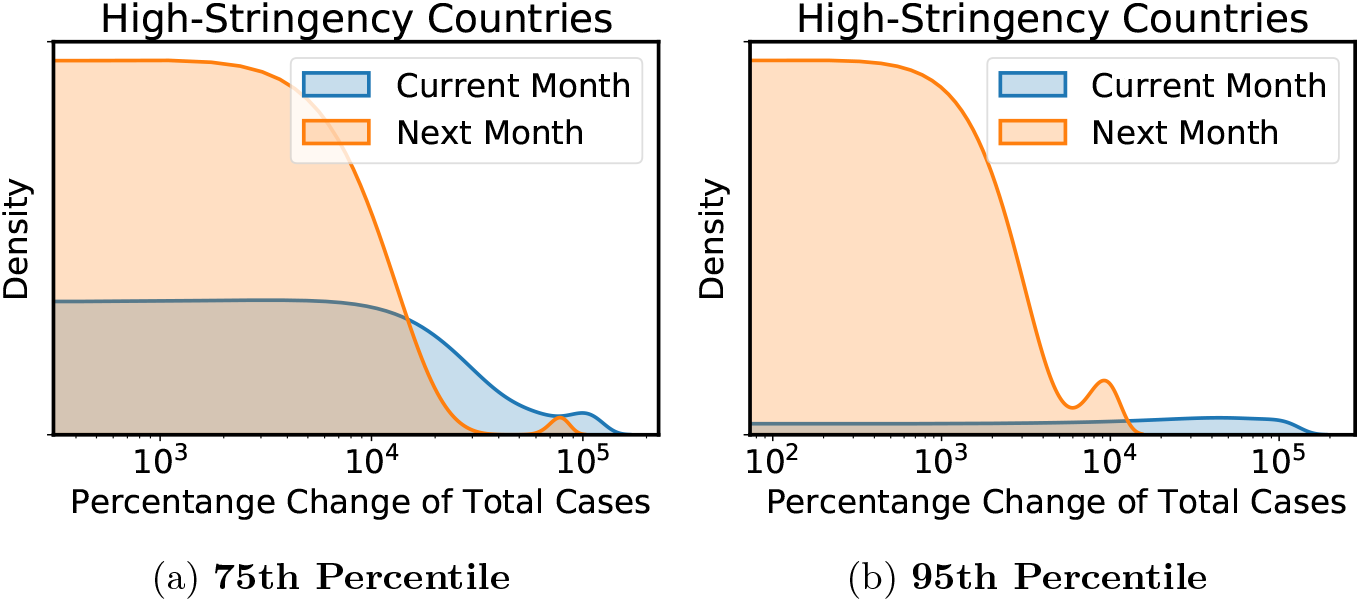
What is the effect of implementing the most aggressive lockdown and restrictive measures? Does it help in reducing the cases? From both 75th and 95th percentiles, it is clear that the area under the curve of the next month has higher concentration on the left side when compared to the current month. This clearly indicates that aggressive lockdown measures have asignificant impact on reducing the number of cases within one month. Using the Spearman Test, percentage change of total cases monthly in the high-stringency countries was analyzed A) from the 75th percentile, the total cases of one month were negatively correlated with the stringency index of the next month (Figure 7 A, r = −0.70, P= 0.0001), and B) from the 75th percentile, there was not significant correlation.

**Figure 8:**
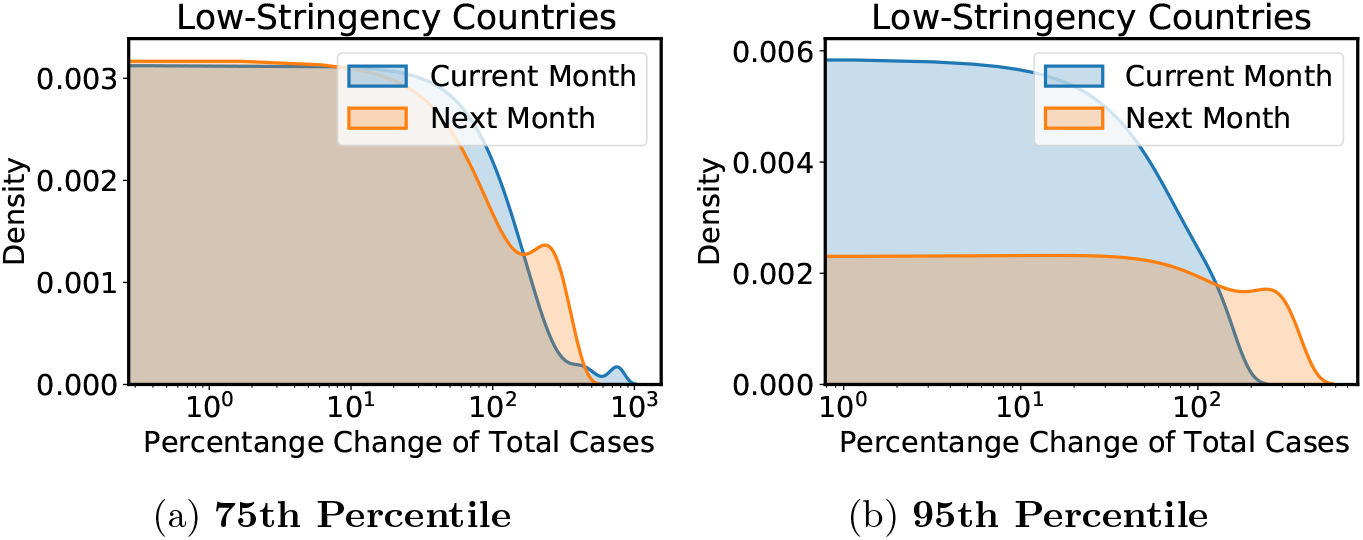
In contrast, what will be the effects if countries have lenient public lockdown measures? Does it have negative consequences? At 75th percentile, the concentration levels are similar for both the current and the next month. However, at 95th percentile, the range of values to the right of the curve for the next month is larger. For the current month, there is a concentration of data on the left meaning that there is a smaller percentage change of total cases when compared to the next month. This clearly shows that lenient restrictive measures have higher tendencies of increased cases after one month. Similarly, the monthly percentage change of total cases in the low-stringency countries was analyzed using the Spearman Test A) from the 75th percentile, and B) from the 95th percentile. There was not significant correlation.

**Figure 9:**
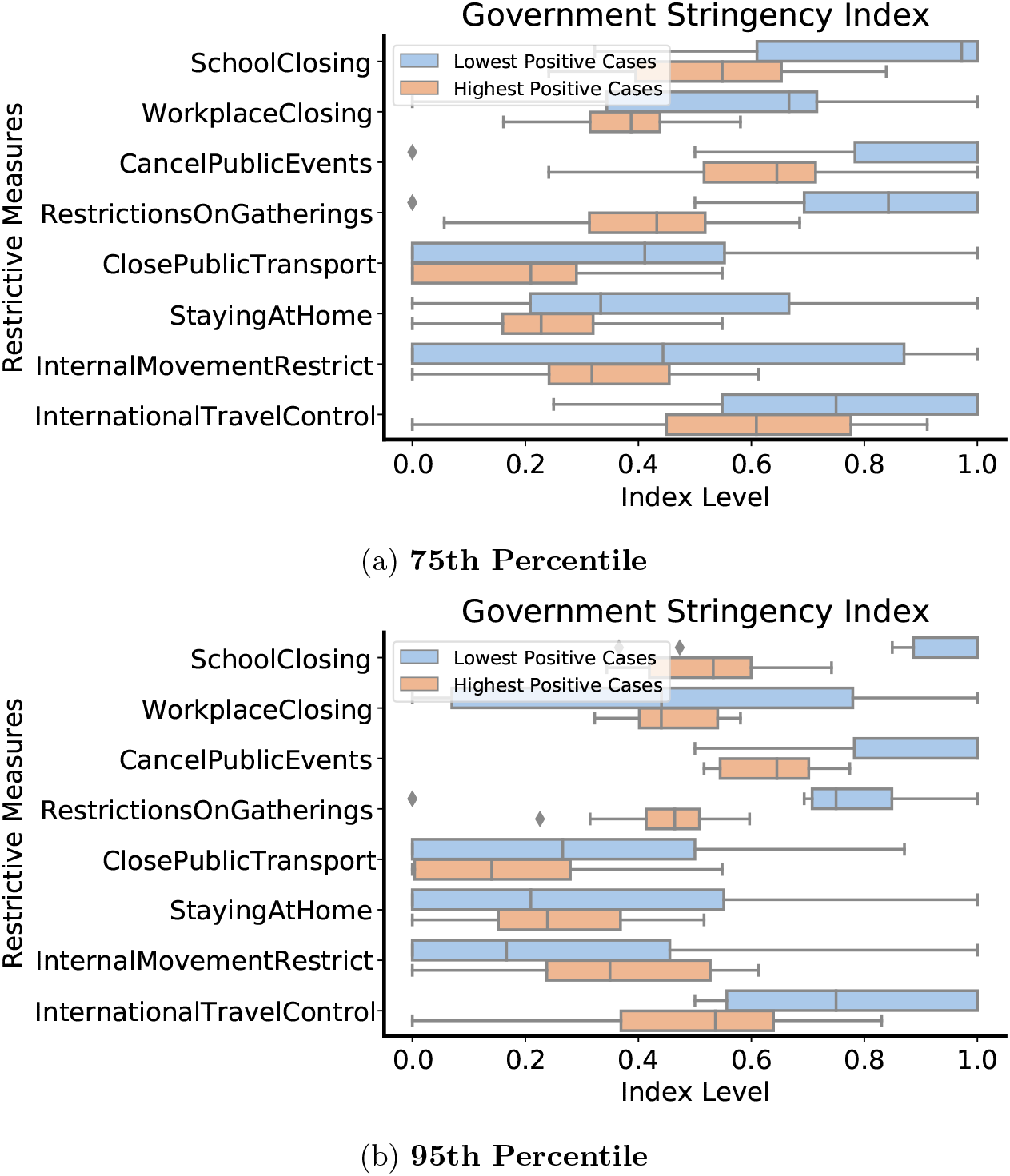
For those countries that were most effective in controlling the COVID-19 outbreak, what steps/measures (i.e., staying at home, internal movement restriction, travel control restrictions, etc.) have they taken that have successfully contributed to outbreak prevention? What are the most important predictor variables that can effectively prevent and minimize the spread of COVID-19? Restrictive measures such as school closing, workplace closing, public event cancellation, restrictions on gatherings, public transport closing, staying at home, internal movement restriction, international travel control in A) 75th percentile countries. There were differences between the countries with the highest or lowest number of positive cases at public event cancellation (P=0.0008), closing public transportation (P=0.03), restriction on gatherings (P*<*0.0001), school closing (P=0.0004), staying at home (P=0.03), and workplace closing (P=0.01). B) 95th percentile countries. There were differences between the countries with the highest or lowest number of positive cases at public event cancellation (P= 0.004), international travel control (P=0.02), and school closing (P=0.001). The data were analyzed using the Student’s t-test.

**Figure 10:**
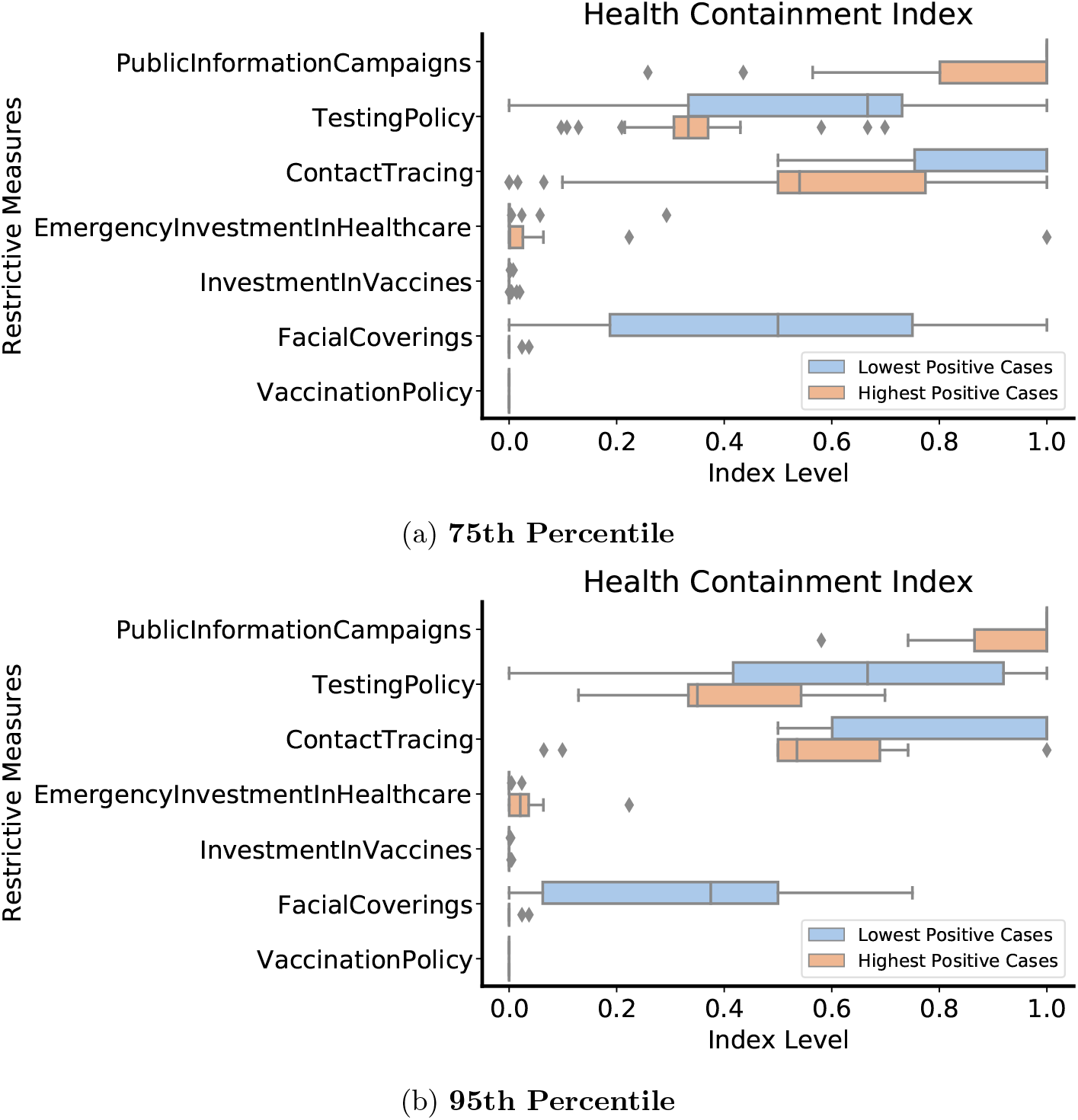
What is the effect of containment measures on COVID-19 outbreak? Containment measures such as public information campaigns, testing policy, contact tracing, emergency investment in healthcare, investment in vaccines, facial coverings, and vaccination policy in A) 75th percentile countries. There were differences between the countries with the highest or lowest number of positive cases at contact tracing (P=0.0006), facial coverings (P*<*0.0001), public information campaigns (P= 0.006), and testing policy (P=0.0002). In B) 95th percentile countries, there were differences between the countries with the highest or lowest number of positive cases at contact tracing (P=0.02) and facial coverings (P=0.002). The data were analyzed using the Student’s t-test.

Finally, we created two groups of countries based on their number of monthly cases: high-case and low-case countries. We then identified the previous month of the stringency index of those countries. This determined if the previous month’s stringency level had any direct impact on the number of cases one month later. We utilized the same method to separate these two groups into the 75th and 95th percentiles, representing high-case countries, and the 5th and 15th percentiles, representing low-case countries (Figure 11).

**Figure 11:**
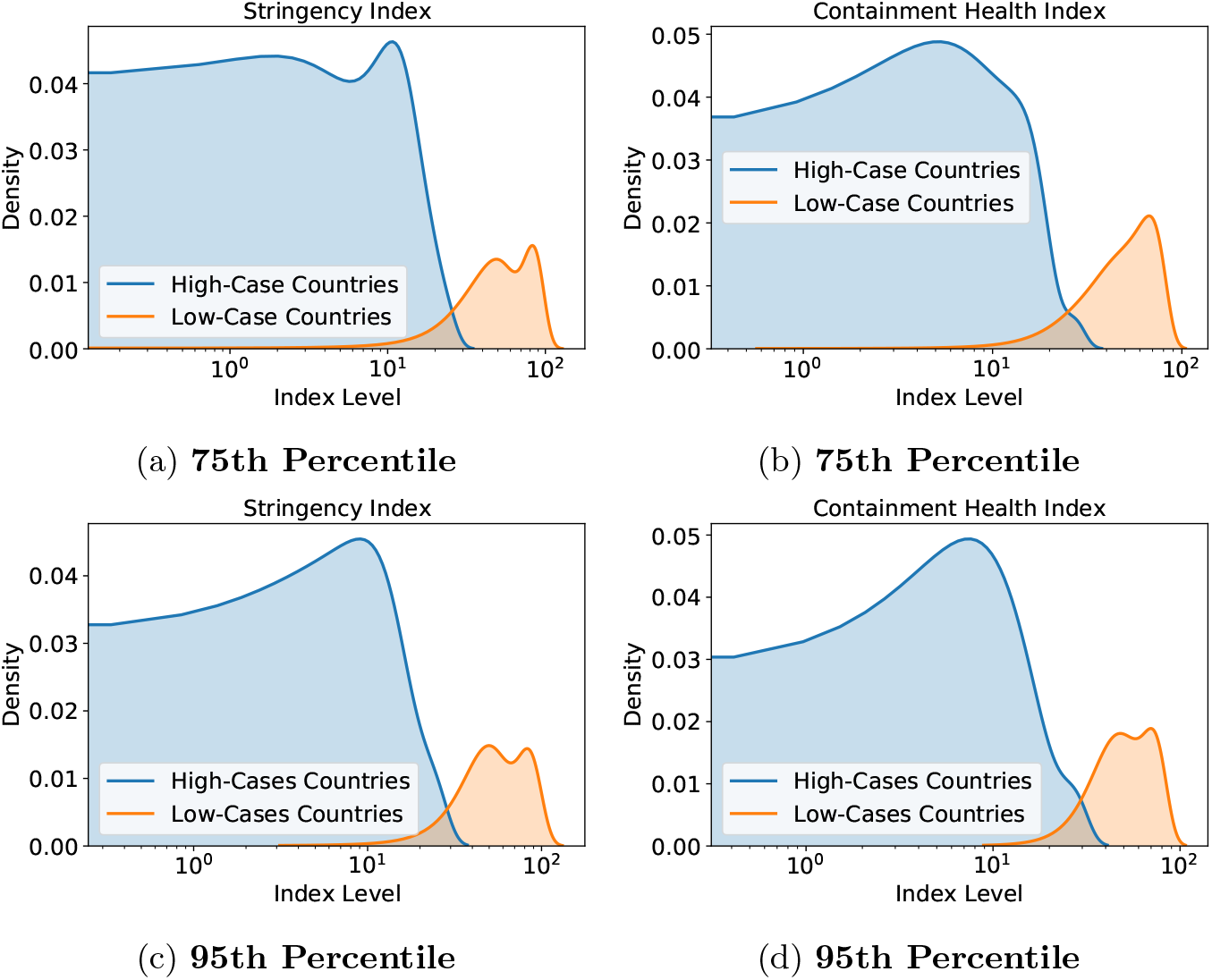
Do countries with the strictest lockdown polices have advantages in containing the virus when compared to countries with lenient lockdown policies? There is a peak around the index level 10 for each graph for the high-case countries. This indicates that the data is concentrated around the index level of 10 for the high-case countries. There is also a peak around the index level 10^2^ for each graph for the low-case countries, which also indicates the data concentration and peak level. Analysis of the difference between low and high cases countries A) stringency index in the 75th percentile countries (P*<*0.0001). B) containment health index in the 75th percentile countries (P*<*0.0001). C) stringency index in the 95th percentile countries (P*<*0.0001). D) containment health index in the 95th percentile countries (P*<*0.0001). The data were analyzed using the Student’s t-test.

We used the Kernel density estimation (KDE) method to create density plots (Figure 1, 4-8, and 11). The KDE is a nonparametric density estimator of the underlying probability density function of the data without assumptions of the population probability distribution functions. The KDE is useful to explore data patterns from a complicated distribution.

Given a set of *N* data points {*x*_1_, *x*_2_, *…, x*_*N*_}, the KDE function 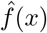 is defined as follows:

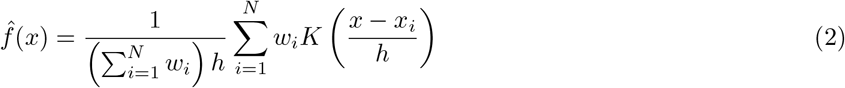

where *x*_*i*_ represents each data point, *K* represents a gaussian function such that ∫ *K*(*x*) *dx* = 1 to provide a smooth estimate (continuous function without discontinuity), *h* represents a controllable bandwidth parameter, and each data point *x*_*i*_ is normalized with a weight 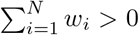 to ensure that 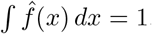. As suggested by [4], the bandwidth parameter *h* is defined as 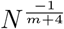 where *m* is the number of features.

For plotting the box plots (Figure 9 and 10), 1.5 is set as the proportion of the inter-quartile range (IQR) past the low and high quartiles to extend the plot whiskers. We identified points outside this range as outliers.

## 3. Results

To assess the effect of population testing, we divided countries into high-testing countries and low-testing countries. The 75th percentile of each group of countries was compared to the total positive cases and the total positive cases per million (Figure 1a and 1b, respectively). Within this sample group, the countries that applied more tests registered a higher number of cases per million (P*<*0.0001, Student’s t-test), but there was no difference between the total number of cases (P=0.13, Student’s t-test). The distribution of cases was similar in the 95th percentile, despite the sample group being smaller (Figure 1c and 1d). Furthermore, we observed that countries with the highest testing had more cases per million (P*<*0.0001, Student’s t-test), with no significant difference in the absolute total of cases when compared to the group of countries with less testing (P=0.18, Student’s t-test).

High-testing countries had no correlation between the number of tests and the number of cases per million. Low-testing countries, however, had a moderate positive correlation between the number of tests and the number of cases per million (r=0.69, P*<*0.0001, Spearman Test). These results demonstrate that high-testing countries have more positive cases per million than low-testing countries. The number of cases, though, is not directly related to the number of tests. In contrast, low-testing countries demonstrate more confirmed positive cases as they perform more tests. These data suggest low-testing countries primarily use tests to confirm cases with symptoms. Thus, a higher positive test rate is a possible outcome. Low-testing countries could consequently have a higher rate of Covid-19 cases than the number of tests confirmed.

To understand improved outcomes of countries with regard to the level of testing, we analyzed the number of new cases and monthly tests in the four countries with the greatest testing (Figure 2). None of these countries showed significant correlation between the number of new cases and the number of monthly tests. In the first months we analyzed, these countries recorded more positive cases than tests per thousand. These data illustrate the countries’ experiences at the beginning of the pandemic. Case numbers exploded, and few tests were available. Following this initial period, Luxembourg, the United Arab Emirates, Denmark, and Cyprus had several months with more testing than positive cases, indicating that these countries were testing the population in a preventive way in order to control of the spread of the virus.

Conversely, countries with low testing tend to have a positive correlation between the number of tests and new cases (Figure 3a-3d). In the Democratic Republic of Congo and Madagascar, the reduction in new cases coincides with the reduction in tests (Figure 3a and 3b, respectively). Moreover, Madagascar had positive correlation between the number of new cases and the number of tests (Figure 3b, r=0.65, P=0.045, Spearman Test). These data reinforce previous data, suggesting that reduced testing availability has limited the detection of new cases in these countries. Countries with high testing also provided reliable data on the spread of the virus. Additionally, we observed a difference between the Case Fatality Ratio (CFR) in the two groups of countries. The 75th percentile showed no difference in the CFR between high-testing countries and low-testing countries (Figure 4a). At the 95th percentile, however, low-testing countries had a higher CFR (mean = 0.0231) than the CFR of high-testing countries (mean = 0.0062, P = 0.003, Student’s t-test, Figure 4b). These results demonstrate that countries that conducted a greater number of tests in a preventive manner decreased the number of deaths and had a lower CFR.

Comparatively, we correlated the CFR with hospital bed capacity, economic output, elderly population, and median population age of the countries (Figure 5). In the 75th percentile, the mean hospital beds per thousand (HBPT) was 8.1 ± 0.6 in the countries with the highest HBPT and was 0.64 ± 0.07 in the countries with the lowest HBPT (P*<*0.0001; 95% confidence interval [CI], −8.8 to −6.2), Figure 5a. In the 95th percentile (Figure 5b), the mean HBPT was 10.6 ± 1.0 in the countries with the highest numbers and the mean was 0.4 ± 0.06 in the countries with the lowest numbers (P*<*0.0001; 95% confidence interval [CI], −12.5 to −8.0). In the 75th percentile (Figure 5 C), the mean GDP per capita was 60738 ± 5329 in the countries with the highest GDP per capita and was 2113 ± 243.9 in the countries with the lowest GDP per capita (P*<*0.0001; 95% confidence interval [CI], −69552 to −47698).

In the 95th percentile, however, the countries with the highest GDP per capita had a mean of 82275 ± 10182, and the countries with the lowest GDP per capita had a mean of 1177 ± 115.3 (Figure 5d, P*<*0.0001; 95% confidence interval [CI], −104579 to −57616). Furthermore, the CFR was negatively correlated with GDP per capita in countries from the 95th percentile (r = −0.64, P = 0.054). We did not observe this in countries of the 75th percentile. These data suggest that the country’s economic situation may impact the CFR. The value of P, though, was borderline (P = 0.05), showing that we cannot confirm a strong tendency in the relationship between CFR and GDP per capita.

Additionally, the mean population aged 70 or older was 14.1 ± 0.4 in the countries with the highest percentage and was 1.4 ± 0.1 in the countries with the lowest percentage (P*<*0.0001; 95% confidence interval [CI], −13.5 to −11.9) in the 75th percentile countries (Figure 5e). In the 95th percentile countries (Figure 5f), the mean population aged 70 or older was 15.7 ± 0.8 in the countries with the highest percentage and was 1.0 ± 0.2 in the countries with the lowest percentage (P*<*0.0001; 95% confidence interval [CI], −16.5 to −12.8). CFR and age also correlated positively in both countries for the 75th and 95th percentile. (r = 0.52, P = 0.003 and r = 0.82, P = 0.006, respectively).

The population difference is more evident in the countries of the 95th percentile, where we confirmed a high positive correlation coefficient between population age and CFR. The mean population age was 44.7 ± 0.5 in the countries with the highest percentage and was 18.7 ± 0.3 in the countries with the lowest percentage (P*<*0.0001; 95% confidence interval [CI], −27.0 to −24.8) in the 75th percentile (Figure 5g). In the 95th percentile countries (Figure 5h), the mean population age was 46.5 ± 0.7 in the countries with the highest percentage and was 17.4 ± 0.3 in the countries with the lowest percentage (P*<*0.0001; 95% confidence interval [CI], −30.8 to −27.3).

We also analyzed if restrictive measures could control the outbreak. We compared the monthly percentage change of total cases between countries with high and low restrictions. In the 75th percentile countries, the mean monthly percentage change of total cases was 66.2 ± 0.7 in the high-stringency countries and was 31.4 ± 1.8 in the low-stringency countries (P*<*0.0001; 95% confidence interval [CI], −38.6 to −31.0), Figure 6a. In the 95th percentile countries, the mean monthly percentage change of total cases was 70.3 ± 1.2 in the high-stringency countries and was 21.4 ± 2.8 in the low-stringency countries (P*<*0.0001; 95% confidence interval [CI], −55.4 to −42.5). This group contained an expressive difference (48.9 ± 3.0) between high-stringency and low-stringency countries.

When comparing the percentage change of total cases month by month, we observed that the total cases of one month negatively correlated with the stringency index of the next month in the 75th percentile countries with the highest monthly jump of stringency (Figure 7a, r = −0.70, P= 0.0001). We did not, however, observe this negative correlation in the 95th percentile countries (Figure 7b). Furthermore, when we analyzed the indexes and cases of the same month, no similar relationship existed. These results suggest that restrictive measures decrease the number of cases, but the decrease only begins after following these measures for at least one month.

In contrast, countries with minor restrictive measures demonstrated no relationship between the measures and the total number of cases. In the 75th percentile, the total number of cases is similar (Figure 8a), and in the 95th percentile we saw differences between the months, but these data were not statistically significant (Figure 8b). It is probably not possible to observe the relationship between the factors as the stringency indexes in these countries remain consistently low throughout the year, while the number of cases varies.

As we observed that countries with the strictest restrictive measures were successful in reducing the number of cases, we analyzed which restrictive measures the countries adopted. The restrictive measures we observed were: school closing, work-place closing, public event cancellation, restrictions on gatherings, public transport closing, staying at home, internal movement restriction, and international travel control (Figure 9). The public event cancellation index was higher in countries with the lowest number of positive cases in the 75th and 95th percentile (P=0.0008 and P= 0.004, respectively), demonstrating that this measure could be important for reducing positive cases.

Moreover, closing public transportation (P=0.03), restriction on gatherings (P*<*0.0001), school closing (P=0.0004), staying at home (P=0.03), and workplace closing (P=0.01) were essential measures for reducing positive cases in the 75th percentile countries (Figure 9a). International travel control and school closing were measures that reduced the number of positive cases in the 95th percentile countries (P=0.02 and P=0.001, respectively, Figure 9b). These results suggest that public event cancellation, international travel control, and school closing were the most important measures to reduce the virus spread.

Likewise, we observed containment measures such as public information campaigns, testing policy, contact tracing, emergency investment in healthcare, investment in vaccines, facial coverings, and vaccination policy in countries with both high and low numbers of positive cases (Figure 10). In the 75th percentile countries, contact tracing (P=0.0006), facial coverings (P*<*0.0001), public information campaigns (P= 0.006), and testing policy (P=0.0002) had elevated index measurements in the countries with the lowest numbers of positive cases (Figure 10a). In the 95th percentile countries, only contact tracing (P=0.02) and facial coverings (P=0.002) were important measures for reducing the number of positive cases (Figure 10b), suggesting these two measures were the most important for controlling the disease. The other measures, however, were also important when considering a large group of countries.

Finally, we found that both the 75th and 95th percentile countries that had the lowest number of cases also had a higher stringency index than countries with the highest number of cases (Figure 11a and 11b, P*<*0.0001, Student’s t-test). The same was observed in relation to the containment health index (Figure 11c and 11d, P*<*0.0001, Student’s t-test), confirming that these measures were essential to control the virus spread and prevent increase in the number of positive cases.

## 4. Discussion

Since the World Health Organization (WHO) declared Covid-19 a pandemic in March, 2020, each country has taken different measures to control the spread of the SARS-CoV-2 virus. To understand the effects of the measures countries have taken to control the Covid-19 outbreak, we analyzed data related to one year of the pandemic. We first observed the effect of the testing level in different countries and the number of positive cases. Countries achieve population testing via the reverse transcription-polymerase chain reaction (RT-PCR), which is the gold standard for detecting SARS-CoV-2 and the only reliable test for determining positive cases [5]. Countries use RT-PCR to control virus spread, especially because asymptomatic or pre-symptomatic people can transmit the virus and infect other people [6]. Thus, countries must perform preventive testing to detect these cases.

We confirmed that high-testing countries had more cases per million than low-testing countries. Only in low-testing countries, however, did the number of tests directly relate to the number of positive cases per million. We confirmed these data when we analyzed information month by month. Even Madagascar, a low-testing country, had a positive correlation between the number of tests and the number of positive cases. Thus, despite high-testing countries having more positive cases during the year, there are months with a low number of new positive cases although the level of testing remained high. High-testing countries could thereby control the spread of the virus. Low-testing countries, however, remained with the number of positive cases similar to the testing level. This suggests these countries may underestimate the number of cases. Consequently, countries with a restrictive testing policy may have total numbers of detected cases that do not correspond to reality.

The study estimated that a large part of the transmission of the virus would occur by people who did not yet have symptoms (35%) and also asymptomatic people, those infected but who did not show symptoms (24%) [7]. Furthermore, a systematic review with data from England and Spain showed that approximately 33% of people who tested positive for COVID-19 were asymptomatic [8]. It would be important for countries to establish criteria to define whether a death was caused by COVID-19. For example, confirming the patient’s contamination with a positive test. Countries that were concerned with correctly diagnosing cases, such as Belgium, had high rates of positive cases for this reason [9]. Understanding the pandemic consequences in countries without this concern, though, is difficult [9].

Moreover, we found that the CFR was lower in high-testing countries, demonstrating that frequent or comprehensive testing, in a preventive manner, led to a decrease in Covid-19 deaths. The CFR was also significantly lower in the 95th percentile high-testing countries than in the 75th percentile, emphasizing that high testing is related to the CFR reduction. In April, 2020, for example, Sweden had a higher CFR than countries that established restrictions such as lockdowns. Furthermore, the UK and France, both low-testing countries, also had a high CFR [10]. Ergonul and collaborators analyzed the CFR of 34 countries and observed that elevated CFR was related to diseases like tuberculosis, disorders such as obesity in adults older than 18 years, and elderly people. On the other hand, the CFR was negatively related to rural population and hospital bed density.

Additionally, the CFR is related to population age and GDP per capita because it is higher in countries with more elderly (aged 70 or older) people and in countries with reduced GDP per capita. The CFR was lower in the countries of the 95th percentile with a low proportion of elderly people than in the countries of the 75th percentile. In Japan, the CFR was 18.1% for people aged 80, 8.5% for people aged 70, and was even lower for people aged 60, with a CFR of 2.7% [9]. Interestingly, the proportion of hospital beds in each country was not related to CFR.

We further analyzed the effect of restrictive measures on the control of the pandemic. We observed that the monthly percentage change of total cases was higher in high-stringency countries, especially in the 95th percentile countries. To this point, we must address the cause-effect relationship individually, as some countries could be more stringent only when the number of cases increased [9]. In the 75th percentile countries, however, the stringency was related to a diminishing number of cases in the following months, demonstrating that restrictive measures are effective in reducing the number of cases, but countries will only realize these effects after four weeks of following these measures. This is an important point of emphasis in awareness campaigns. The population must be aware that it takes at least four weeks for the restrictions to reflect the number of new cases. In countries with lenient public lockdown measures, we observed no significance regarding the change in the number of cases. This was likely due to the lack of measures to decrease the number of cases. Thus, the number of new cases remained stable.

We observed that both the 75th and 95th percentile countries with a reduced number of cases had higher stringency indexes. Our work verified that public event cancellation, international travel control, school closing, contact tracing, and facial covering were the measures related to the reduction of cases. In any type of event, distancing between people is the most efficient way to avoid contamination [11]. During outbreaks, leaders and citizens must prevent the occurrence of public events and avoid the contact of people who are not from the same family. In situations with many people in close proximity, such as public transport, it is difficult to estimate the efficacy of individual protection measures, such as masks, as the exposure is high [11]. In Japan, prohibition of sporting events, concerts, and school reduced the R0 from 2.5 to 1.1, and after reopening, the index was higher than the initial one [12]. This indicates that prohibition of events is effective in reducing the number of new cases. Our results corroborate these data.

School closing was also a measure that countries with a low number of cases applied. In the USA, between March and May, 2020, school closing led to a decrease in the number of cases and mortality [13]. A systematic review in April, 2020, showed that the school closures did not impact the decrease in the transmission of the SARS-CoV-2 virus [14]. We, however, analyzed data from a year of the pandemic, making it possible to assess the emergence and control of outbreaks in several countries. Either way, schools should have restrictions, such as the use of masks and distancing between students, to avoid contamination [15].

The World Health Organization recommends the use of facial coverings (surgical masks, N95, or cloth face coverings) to prevent the spread of the virus. Evidence of the efficacy of the use of facial coverings exists, such as a case in the USA in which two hairstylists worked for a few days while infected with the SARS-CoV-2 virus. Due to the use of masks, however, the hairstylists contaminated none of the 139 clients they serviced [16]. Another study looked at data from 200 countries, and facial coverings were effective in reducing the number of infections and deaths [17]. People must, however, adjust masks to fit well so the mask filters all of the air exchanged [18]. These data are in agreement with our results, showing that the use of facial coverings was an essential measure for the reduction of cases.

Research has shown social isolation and case tracking are effective in controlling the spread of the virus if done properly [19]. The risk of contamination is higher when there is more exposure to the virus. Ideally people keep a distance of 2 m and limit contact time to less than 10 minutes with an infected person [20]. Contact tracing will only be effective if people observe short contact times with each other [21]. An infected person can have no symptoms and still transmit the virus to another person. With case tracking, it is possible to alert someone that had close contact with an infected person, even if it is someone who has had contact in a public environment. Moreover, mobile apps for contact tracing exist, and they improve the process with rapid and anonymous identification [20]. Apps must prioritize detection efficiency and user privacy [22]. A survey in Belgium showed that only 49% of people would use a tracing app. The main concern was privacy [23]. However, this strategy will only be successful if it is a comprehensive screening approach and the tracked people obey adequate isolation [24][21]. Our results showed that contact tracing was important to prevent the virus from spreading during the outbreak.

In China, screening for positive cases was more efficient in containing the spread of the virus than international travel control [25]. A study that evaluated several countries, however, showed that restricting international travel was important to reduce mortality [26]. Our data demonstrate that international travel control was an important measure for the control of the pandemic during the year 2020. Countries should continue to consider restricting international travel control for the current year, primarily to avoid the contagion with new emerging mutations in some countries.

Finally, it was observed that both the 75th and 95th percentile countries with higher stringency indexes had reduced numbers of cases. Furthermore, restrictive measures affect the number of new cases after four weeks. The measures related to the reduction of the cases were public event cancellation, international travel control, school closing, contact tracing, and facial covering. These results will be essential to the understanding of a year of pandemic, especially for countries that still face the outbreak. Despite scientific advances about the Covid-19 disease and the beginning of vaccination, many countries still face a critical situation and the emergence of new variants is a warning. Overall, restrictive measures are necessary to contain the spread of the virus and, consequently, reduce mortality.

## 5. Conclusion

In this paper, we have performed a comprehensive examination of the long-term effects of the COVID-19 pandemic using one year of public data. We also evaluated several government intervention variables and restrictive measures. We found evidence that restrictive measures reduce the number of new cases after four weeks, indicating the minimum time required for the measures to be established. We also found that public event cancellation, international travel control, school closing, contact tracing, and facial coverings were the most important measures to reduce the virus spread. Moreover, we observed that countries with the lowest number of cases had a higher stringency index.

Our findings are relevant for decision-makers in implementing appropriate intervention policies. Since countries have not widely adopted COVID-19 vaccination during the first year of the outbreak, we did not have sufficient data to evaluate the effectiveness of vaccine adoption. Therefore, future research work may examine the short-term and long-term effects of different vaccination strategies. Finally, we may also utilize publicly available mobility datasets to assess the joint effects of government interventions, social distancing, and economic policies with respect to population movements.

## Data Availability

Data available upon request.

